# The effect of peer support networks in alleviating anxiety and enhancing perceived social support

**DOI:** 10.64898/2026.03.22.26349028

**Authors:** Wan-Jou She, Benjamin Yip, Alexandra Covaci, Shibuya Yu, Chee Siang Ang, Shinsuke Nakajima, Panote Siriaraya

## Abstract

Support from peers has long been considered an alternative support resource than professional healthcare ones. Despite the inconclusive findings of previous studies regarding the effects of peer support, the integration of Peer Support Networks (PSNs) for youth and adolescents appears to offer promising outcomes. However, many existing digital peer support systems operate as proprietary platforms, lacking transparency in monitoring the efficacy of support and in understanding how personality traits influence outcomes.

However, many existing digital peer support systems operate as proprietary platforms lacking transparency in monitoring the effect of peer support and understand the influence of personality traits on its outcomes. To address these limitations, we utilized our research platform, Peer2S, a digital PSN designed to facilitate connections based on shared lived experiences while simultaneously monitoring users’ mental well-being and personality traits.

We conducted a four-week within-subjects experiment with 28 Japanese university students to examine the PSN system’s impact on anxiety and perceived social support. Following a two-week baseline control period, participants interacted with the system for two weeks. Pre- and post-intervention assessments utilized generalized anxiety and multidimensional social support measures, alongside personality evaluations. The results indicated that participants experienced a significant reduction in anxiety after using the system, whereas no significant changes occurred during the control period. Perceived general social support showed a borderline significant increase, though specific college-context support dimensions remained unchanged.

Furthermore, multiple regression analysis revealed that personality traits moderated anxiety outcomes. Contrary to typical protective associations, higher agreeableness significantly predicted increased anxiety during the intervention, which may reflect cultural tendencies toward conflict avoidance and over-accommodation in Japan. Conscientiousness demonstrated a marginally significant protective effect against anxiety, while personality traits did not predict changes in perceived social support.

These findings suggest that short-term, algorithmically mediated peer support can yield measurable improvements in mental well-being, particularly in reducing anxiety. Moreover, the varying impacts of personality traits highlight the necessity of considering sociocultural contexts when designing and deploying digital mental health interventions.

**Author’s summary:** The formation of social bonds is often selective, established through shared values, cultural interests, or significant life experiences among “peers.” In some populations such as adolescents and young adults, peer support is regarded as a promising source of empathy, understanding, and psychological support.

We report a study conducted using our customized digital peer matchmaking system with Japanese university students to examine if this novel approach to peer support impacts mental well-being. We found that after just two weeks of using the system, participants experienced a significant reduction in their anxiety levels. We also dove deeper to look at if individual personality traits influence their use outcomes. Interestingly, our results revealed that highly agreeable individuals actually experienced increased anxiety while using the system. In a Japanese cultural context, this may occur because agreeable users tend to avoid conflict and over-accommodate others at their own expense. Ultimately, our research demonstrates that matchmaking algorithms can effectively facilitate digital peer support to improve mental well-being, provided we carefully consider how different personality traits and cultural backgrounds shape user experiences.

## 1. Introduction

As inherently social beings, humans naturally seek connection, recognition, and support from those around them. Social circles are often formed and reinforced through shared experiences, cultural interests, or significant life events that foster deep empathy and strengthen bonds (1). Among these connections, individuals who share similar experiences, circumstances, or challenges are often referred to as peers (1,2).

Having experienced comparable struggles, such as anxiety, depression, trauma, or loneliness, peers play a crucial role in providing understanding, validation, and a unique sense of connection. More importantly, the ability to connect with peers can help alleviate feelings of isolation and loneliness for those navigating hardship (3–7). With these potential benefits for mental health, and driven by the growing demands, Peer Support Networks (PSNs) have increasingly become an integral part of the healthcare system, complementing formal healthcare resources by offering unique benefits derived from lived experiences (3–6,8–10).

Although the evidence regarding the impact of PSNs in healthcare remains inconclusive (partly because peers may lack the professional expertise required for effective assistance), certain populations such as youth and adolescents and older adults have emerged as groups that benefit significantly from PSNs (4). A review paper by Shalaby and Agyapong reported that youth and adolescents are a key demographic “particularly in need” of peer support due to widely documented challenges such as social isolation and loneliness (4). In addition, peer supporters for adolescents are seen as a more effective yet with minimal adverse outcomes (11) alternative to untrained or inexperienced close friends or unfamiliar professionals (7).

Paradoxically, merely offering a “platform” to connect peers may not suffice. Modern prevalence of social media use, a service that could readily incorporate PSN features, among young populations is widely criticized for exacerbating youth mental health problems such as loneliness and anxiety (12,13). For instance, the well-documented negative impact such as toxic comparison culture fueled by the number of likes (14–16), unrealistic portray of lifestyles through reels (17) and cyberbullying caused by anonymous public comments (18,19).

On the other hand, many studies have found positive effects when such platforms are specifically designed to support PSN (9,10,20), some even utilizing platforms like Facebook and Instagram (21–24). Reported benefits include an improved sense of normalcy (25,26), expanded social networks (25,27), increased satisfaction with peer interactions (28), reduced isolation (27), a stronger sense of community (27), and greater self-efficacy and agency (29).

Nevertheless, there remains limited evidence and understanding to identify the underlying mechanisms through which such peer support exerts its benefits and supports youth and adolescents with mental health concerns. As most existing PSNs operate as proprietary systems, there is a need for a more transparent PSN design to provide clarification regarding the support process and to discern the possible elements that contribute to its effectiveness.

As such, we developed and implemented one such PSN system (Peer2S) centered around sharing of lived experiences and mutual supporting (30). To reduce the risk of comparison or pressure that are often caused by SNS platforms publicity, the Peer2S system limits reactions’ visibility to the users themselves (e.g., number of support messages and hugs). The peers’ stories are matched and recommended using the algorithm similar to the commercial-style matchmaking app, based on individuals’ demographic background, interests, concerns and lived experiences.

We conducted an experiment study to examine and better understand the impact of a PSN on college students’ mental health, specifically on anxiety and social support. According to the above discussion, we come up with the following hypotheses:

H1: After using the Peer2S system for 2 weeks, participants would experience a significant increase in perceived social support, while no meaningful change is expected over the preceding 2-week control period.

H2: After using the Peer2S system for 2 weeks, participants would experience a significant decrease in anxiety, while no meaningful change is expected over the preceding 2-week control period.

H3: Participants’ personalities will have a significant impact on the changes on perceived social support and anxiety level when using the Peer2S system.

## 2. Method

To evaluate the effectiveness of our peer-based matchmaking approach, a within-subjects experiment was conducted with 23 university students from Japan, in which participants used the Peer2S system for two weeks and pre–post changes in measures such as anxiety and perceived social support were evaluated relative to a preceding two-week baseline period. The study method has been reviewed and approved by the institutional review board (IRB) at a private university in Japan (Kyoto Sangyo University IRB No:0159). The detailed methods are explained in the subsections.

### 2.1 Participants

Study participants were recruited within the campus through flyers and word of mouth reference. The inclusion criteria are students who: (1) are aged 18 to 24, (2) are actively enrolled in the university, (3) speak Japanese, and (4) have internet access which can be used on either a computer or mobile device. Participants who indicated interest to participate in the study would sign the Informed Consent Form and proceed to register their profiles in the Peer2S system.

### 2.2 Study Design

Before the experiment was carried out, all participants were asked to register on their profile on the system, regardless of their order of enrollment. We did this to ensure that the system had enough profiles to recommend to each participant. The main study lasted for four weeks. At the beginning (first checkpoint) and at the end of the first two weeks (second checkpoint), participants completed all psychological measures (see Section 3.2 for introduction to the measures we used) to establish baseline levels of anxiety and their social support (first checkpoint). These first two weeks served as the control period. In the second phase of the study, which lasted for the following two weeks, participants were asked to interact with the application. After the second checkpoint, participants would begin using the Peer2S system for two weeks.

During the 2-week study, each participant received recommended profile from five potential peers daily. Participants were able to review the profiles and decide whether and which peer(s) to match or interact with (e.g. by sending supportive messages or giving virtual hugs). If both sides agreed to match with each other, the conversation function was enabled, allowing participants to chat with each other during the duration of the study. The Peer2S system collected their match decisions and chat logs for further analysis. At the end of the two-week matchmaking period, we asked participants to complete the final set of measurements, including user experience assessments and the same psychological measures (third checkpoint)

### 2.3 Measurements

#### 2.3.1 Generalized Anxiety Disorder-7 (GAD-7) Japanese Version

The GAD-7 was developed as a self-report questionnaire to assess generalized anxiety and consists of seven items (31). These items include factors such as whether the respondent feels tense or irritable, whether they find it difficult to relax, and whether they feel they are frequently worried. The Japanese version which was developed by Muramatsu (32) was used in this study.

#### 2.3.2 Multidimensional Scale of Perceived Social Support (MSPSS)

We use the Japanese version of the Social Support Scale (Multidimensional Scale of Perceived Social Support) developed by Iwasa et al. (33), based on the original version by Zimet et al. (34). This scale measures the degree of social support and connectedness that subjects perceive from others. It has the same three-factor structure as the original version, namely, family support, significant other support, and friend support. In this study, we only use the friend support subscale, redefining friends as “supporters found through the application.”

#### 2.3.3 Social Support Scale for College Students (SSSCS) Japanese Version

To obtain a more nuanced understanding of the Japanese college students’ social support status, we also used the social support scale developed by Japanese researchers (片受靖 & 大貫尚子, 2014). The questionnaire contains 23 items answered on a 4-point Likert scale. The scale is divided into three factors: appraisal support (e.g., recognition and acknowledgement from others), informational / instrumental support and emotional / companion support. The differences between this scale (SSSCS) and the one used to measure perceived social support (MSPSS) is summarized in Table 1 for clarity.

**Table 1.** Key differences between the two social support scales.

| **Feature** | **MSPSS (Friend Support)** | **SSSCS (College Students Support)** |
| --- | --- | --- |
| **Focus** | Perceived support from friends | Various types of social support specific to the context of Japanese college students |
| **Structure** | One subscale (friend support) from a larger 3-factor model | 23 items divided into 3 detailed support categories |
| **Measurement** | How much support students feel they get from their app-based social network | More detailed breakdown of the specific types of support received from their peers |
| **Context** | Based on Zimet et al. (1988), adapted for Japan | Designed specifically for Japanese students |

#### 2.3.4 Big 5 Personality Survey

We assessed participants’ personality traits using the Big5 Personality Survey, a widely recognized instrument in psychological research, also known as the Five-Factor Model (FFM). Originally developed by McCrae and John(36), this survey measures five fundamental dimensions of personality: Openness to Experiences, Conscientiousness, Extraversion, Agreeableness, and Neuroticism. For our study, we employed a validated version that utilizes a 5-point Likert scale. This instrument provides valuable insights into individual differences, helping us explore how personality traits may influence technology-mediated peer-based support obtained through the Peer2S system.

### 2.4 Data Analysis

After checking for missing data, measurement errors and unusual values from the questionnaires and a clean dataset was prepared for the quantitative analysis. The data from surveys described in Section 4.2 were entered into SPSS to perform the one-way repeated measures ANOVA, or the Friedman test if the data were non-normally distributed. Since our primary research questions focused on examining whether there were changes in anxiety and perceived social support during the control period (e.g. from “2 weeks before” to “Beginning of PEER2S session”) and after using the system (from “Beginning of PEER2S session” to “End of PEER2S session”), only these two pairwise comparisons were tested. Accordingly, we controlled the familywise error rate using a Bonferroni correction over these two post hoc comparisons. Additionally, Multiple regression analysis was conducted to assess the relationship between the personality traits of users and the effectiveness of the system on anxiety and perceived social support.

## 3. Results

A total of 28 students participated (15 males and 10 females, 3 preferred not to disclose). Interaction data was available for 23 of the participants. In total, 50 chatrooms were created (i.e., 50 matched pairs), and 275 messages were sent. Additionally, 17 chatrooms were created but never used (66% active chatrooms), and one user never matched with any other peer (97.92% match rate). Regarding the interaction patterns, 82% of the participants sent at least one hug (Mean = 9.32, SD=7.74). All 23 participants received at least one hug (Mean=7.78, SD=3.30). With regards to support messages, 73.91% of participants sent at least one support message (Mean=3.70, SD=4.04) and 95.6% of participants received at least one support message (Mean=2.63, SD=1.32).

**Fig 1:**
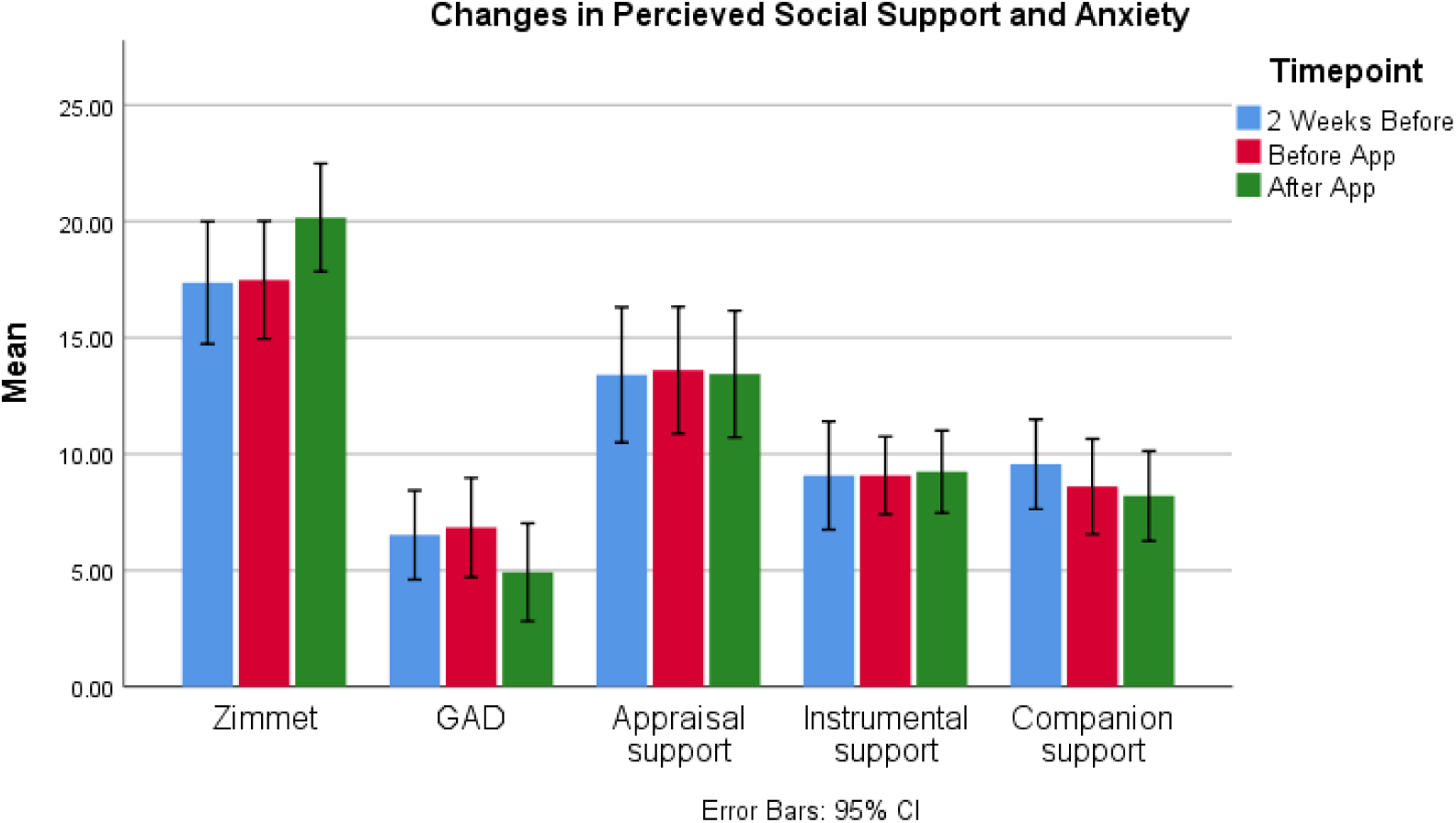
Changes in measures related to mental wellbeing.

### 3.1 Potential Increase in Perceived Social Support Following PEER2S Session

After confirming that the assumptions of normality and sphericity were met and verifying the absence of outliers, a one-way repeated measures ANOVA was conducted to evaluate the effect of the 4-week PEER2S intervention on scores from the Multidimensional Scale of Perceived Social Support (Zimet) in the sample of 28 participants.

Assuming sphericity, the repeated measures ANOVA revealed a statistically significant effect of the PEER2S intervention on Zimet scores over time, F (2, 48) = 4.181, p = 0.021. Post hoc comparisons with Bonferroni correction showed no significant change in mean Zimet scores from “2 weeks before” (M = 17.36, SD = 6.38) to the “Beginning of PEER2S session” of the study (M = 17.48, SD = 6.12) (p = 1.000) (e.g. the control period). However, the increase in perceived social support from the “Beginning of PEER2S session” to the “End of PEER2S session” (M = 20.16, SD = 5.63) showed a borderline significant increase (p=0.058).

### 3.2 No Significant Changes in College Social Support Following PEER2S Session

Given that all three dimensions of the Social Support Scale for College Students were not normally distributed, the non-parametric Friedman test was used to determine whether there were significant differences across the different timepoints. The results showed that there was not a significant difference in appraisal support (p=0.83), informational & instrumental support (p=0.623) and emotional & companion support (p=0.918) between the different time points.

### 3.3 Small but Significant Reduction in Anxiety Scores Following PEER2S Session

Given that anxiety was not normally distributed the non-parametric Friedman test was utilized to determine if there were differences in GAD-7 scores. The Friedman test revealed statistically significant differences in GAD-7 scores across the time points, χ²(2, N = 25) = 7.103, p = 0.029. To measure the effect size, Kendall’s W was calculated, yielding W = 0.142, indicating a small effect size. Pairwise comparisons were performed with a Bonferroni correction for multiple comparisons. The post hoc analysis showed a significant differences between the “Beginning of PEER2S session” (Median = 7) and “After PEER2S session” (Median = 3), p < 0.05, while there was no significant differences in the anxiety scores from “2 weeks before” (Median= 5) and in “Beginning of PEER2S session” of the study (Median= 7)(p=1.000).

### 3.4 The Impact of Personality Traits on Anxiety and Perceived Support

Multiple regression analysis was carried out to evaluate the impact of personality traits on change in anxiety and perceived support scores throughout the study. Previous literature has described an interlinked relationship between personality, anxiety and perceived support (37–40). From this analysis, we hope to determine whether individuals with certain personality traits may derive greater or lesser benefit from a matchmaking-based digital peer support system.

#### 3.4.1 Agreeableness Predicts Anxiety; Conscientiousness Approaches Significance

The assumption tests indicated that the residuals were independent (Durbin-Watson = 2.055), with no violations of linearity or homoscedasticity, and no signs of problematic multicollinearity (acceptable VIF and tolerance values). Overall, the model explained 45.2% of the variance in GAD scores (adjusted R² = 29.1%) and was significant (p < 0.05). Among the Big Five traits, Agreeableness was a significant predictor (B = 0.516, p = 0.010), indicating that higher agreeableness was linked to a greater increase in GAD change scores (hence, greater anxiety). Conscientiousness showed a borderline countering effect in GAD change scores (B = −0.453, p = 0.060), whereas Extraversion, Neuroticism, and Openness were not significant predictors.

#### 3.4.2 Personality Traits Show No Significant Impact on Perceived Social Support Changes

The assumption tests indicated that the residuals were independent, with no signs of problematic multicollinearity or influential outliners. The overall model was not statistically significant (p = 0.919) and explained just 7.5% of the variance in ZIMET score changes (adjusted R² = −19.6%). None of the personality traits, including Extraversion, Agreeableness, Conscientiousness, Neuroticism and Openness, significantly predicted changes in perceived social support. Overall, the results suggest no influence of personality traits on ZIMET score outcomes.

Similarly, none of the Big Five personality traits (Extraversion, Agreeableness, Conscientiousness, Neuroticism, Openness) emerged as significant predictors for changes in the scores of all three types of college social support (appraisal support, informational & instrumental support, and emotional & companion support). The assumption tests indicated that the residuals were independent, with no signs of problematic multicollinearity or influential outliners. For appraisal support, although the model accounts for roughly 20.6% of the variance in appraisal support (R² = 0.28), the overall regression was not statistically significant (p = 0.515), and none of the individual traits yielded p-values below 0.05. Similarly, the models for informational/instrumental support (R² = 0.077, p = 0.916) and emotional/companion support (R² = 0.103, p = 0.848) were also nonsignificant. These results suggest that personality traits do not reliably predict changes in any of the measured dimensions of perceived social support.

## 4. Discussion

### 4.1 The Effect of PEER2S on Social Support

Overall, our results suggested that after using the PEER2S system for a period of two weeks, participants showed a borderline significant increase in perceived social support based on the ZIMET score. Furthermore, no changes were observed during the preceding two-week control period, indicating that the observed increase was more likely attributable to the PEER2S system rather than natural variation over time.

However, our results also suggested that usage of the PEER2S system did not produce statistically significant changes within the three dimensions as measured by the social support scale for Japanese College Students. Therefore, H1 was only partially supported.

Such a finding aligned with previous studies’ that claimed sharing of lived experiences through peer interaction could facilitate trust and meaningful relationship (41) and reduce feelings of isolation and loneliness (42). However, since PEER2S system was designed with the very intention to facilitate connections through lived experiences sharing, it should not be surprising to the researchers that the activities involved offering deeper level of support (e.g., instrumental support or companion support, as measured by SSSCS) were not directly mediated by PEER2S with its current design.

Nevertheless, it is worth considering whether such “lip service” social support can produce any meaningful benefits for individuals. From Granovetter’s weak ties perspective, the single-purpose nature of these connections (empathizing through shared lived experiences) and the sense of inclusion in community they offer are considered equally beneficial and socially capitalizable as those derived from “strong ties” (43). They simply serve different functions. Typical examples include kinship versus mentor–mentee relationships, or close friendships versus professional relationships. Therefore, it became clear to us that most connections facilitated by PEER2S are likely to be purpose-driven, peer-support-based weak ties initially; however, this does not diminish their value and the versatile and immediate comfort they could offer to cumulatively brighten our day-to-day lives.

Moreover, a few of these connections may eventually develop into strong ties as the level of self-disclosure increases. As Altman and Taylor’s Social Penetration Theory (often used with an onion metaphor) offers a clear framework regarding how weak ties are established and nurtured into strong ties. Their 4-stage relationship is linear and progressive, meaning strong ties cannot be established by leaping through each stage, and weak ties are always necessary first steps to nurture them (44,45).

Clearly, the current PEER2S has demonstrated good potential in facilitating the first two stages of exchanges. Comparing to befriending on Facebook, users matching on PEER2S would already be considered met the desire to move into the second stage, explorative affective exchange (46). The promising next step will be linking this results to more in-depth disclosure and tangible social support suggested by SSSCS. Moreover, the multimodal social exchanges (e.g., from PEER2S to email or other SNS) are also observed in our study and will require more investigations.

### 4.2 The Effect of PEER2S on Anxiety Reduction

Our results showed that participants exhibited a significant reduction in anxiety following two weeks of using PEER2S, while no changes were observed during the preceding two-week control period, suggesting that the observed reduction in anxiety could be attributable to the intervention rather than time effects. Such a finding aligns with our pilot study results (30), suggesting a *sustainable* and *replicable* effect of using PEER2S. Accordingly, H2 was supported.

Several mechanisms can explain the reduction in anxiety during the intervention. One possible explanation may be the emotional and social support facilitated by peer support interactions. This is evidenced by Richard’s et al. (42) review, which found that peer support inherently fosters a sense of connection, reducing feelings of isolation and loneliness that often exacerbate anxiety. By providing a shared space where participants felt understood and valued, the intervention likely alleviated these feelings, contributing to the decrease in anxiety scores.

In addition, the sharing of coping mechanisms through peer support interactions may contribute to reduced anxiety levels. This aligns with Egmose’s et al. (47) meta-analysis which found that with regular interaction and the exchange of strategies, individuals developed more effective ways to manage stress and adversity. This improvement in coping capacity may have contributed to the observed reduction in anxiety symptoms, as resilience and adaptive strategies play a vital role in anxiety management.

Furthermore, sharing coping mechanisms and lived experiences provides positive role modeling, reinforcing the impact of peer support in alleviating anxiety. Peer supporters who shared their experiences and demonstrated successful anxiety management provided tangible examples of recovery and hope (48). This modeling effect likely inspired participants, showing them that progress is achievable and motivating them to persevere in their own journeys toward anxiety reduction.

Moreover, Egmose et al. (47) highlights that peer support has a positive impact on self-esteem and fosters a sense of empowerment, both of which are crucial for alleviating anxiety. Being part of a supportive peer group may have helped individuals recognize their strengths and abilities, boosting their confidence and self-worth. This increased sense of empowerment is crucial in the context of anxiety reduction, as it fosters a greater sense of control and agency in managing one’s mental health (47). Although the post hoc analysis with Bonferroni correction yielded a borderline significant change, the overall trend indicates meaningful psychological benefits that should not be overlooked.

Lastly, the sense of community and belonging fostered by peer support is another crucial element. Being part of a network of individuals with shared experiences created a supportive environment where participants felt understood and accepted (42). This sense of belonging has been shown to mitigate anxiety by reinforcing the idea that individuals are not alone in their struggles. Therefore, the community aspect of the intervention may have played a significant role in the reduction in anxiety observed.

### 4.3 Evaluating impact of Personality Trait on Anxiety and Perceived Social Support Scores

Our findings indicated that among Japanese college students, those who were more agreeable were associated with higher anxiety scores, whereas higher conscientiousness was borderline associated with lower anxiety scores. While our findings regarding conscientiousness were aligned with previous research (49–51), the inconclusive findings related to anxiety is particularly interesting when placed in a Japanese society context.

Agreeableness interpreted as conflict avoidance at the cost of over-accommodation.

Findings on the association between agreeableness and anxiety are mixed. A few studies on Chinese young adults found strong evidence of agreeableness as a protective factor against anxiety (49,52,53), consistent with the view that prosociality, cooperativeness, and trusting are generally associated with better mental health outcomes and lower anxiety risk (54). However, other studies indicated weak relationship (55) or positive association between agreeableness and anxiety (56). Such a mixed finding suggests interpersonal orientation is not uniformly protective across contexts. In Japanese cultural settings where social harmony and relationship maintenance are highly valued (57), agreeableness may be expressed as conflict avoidance and over-accommodation, which can delay direct problem solving or require suppression of personal needs (58). Such avoidant interpersonal coping is consistent with evidence linking agreeableness to an avoiding conflict style, and may help explain why higher agreeableness predicted less favorable anxiety change in the present study.

Another possible interpretation concerns the distinct roles agreeableness may play in cross-sectional versus pre-post mental change. While higher agreeableness is often associated with lower anxiety in cross-sectional studies (49,52,53), likely reflecting its effect through harmony maintenance, social acceptance, and reduced conflict, it may not translate into equal symptom improvement in a longer-term period. Specifically, agreeableness-linked tendencies toward accommodation, indirect communication, and self-silencing can provide immediate interpersonal relief but may simultaneously impede core therapeutic processes such as assertive boundary-setting, emotional expression of distress, or direct confrontation of maladaptive beliefs (58,59). Over the two-week intervention window, these dynamics could constrain symptom reduction: individuals with conflict-avoidant and self-silencing tendencies limit engagement with core therapeutic processes (e.g., emotional expression, cognitive reframing), even while preserving short-term interpersonal stability. This pattern reflects a well-documented social phenomenon in Japan, where high agreeableness (particularly expressed as conflict avoidance and over-accommodation) can instead trigger higher anxiety.

### 4.4 Conclusion

This study examined the psychological impact of a two-week technology-mediated Peer Support Network (Peer2S) intervention among 28 Japanese college students, focusing on anxiety and perceived social support. Consistent with our hypotheses, the intervention was associated with a significant reduction in anxiety relative to the preceding control period, replicating findings from our pilot study and suggesting that short-term, algorithmically mediated peer support can produce measurable improvements in mental well-being. Perceived social support demonstrated a positive trend following system use, although changes did not reach conventional levels of statistical significance across all measures. Together, these findings indicate that even brief exposure to structured, lived-experience-based peer network may offer meaningful psychological benefits.

Beyond intervention effects, our findings further contribute to the literature by examining the moderating role of personality traits. Notably, higher agreeableness predicted greater increases in anxiety scores across the study period, a pattern that diverges from much of the existing literature positioning agreeableness as protective. Within the Japanese sociocultural context, where interpersonal harmony and conflict avoidance are highly valued, agreeableness may manifest as over-accommodation or self-silencing, potentially limiting assertive coping and amplifying internal distress. Conscientiousness, in contrast, demonstrated a marginal protective effect, aligning with prior findings linking self-regulation and goal-directed behavior to reduced anxiety.

However, personality traits did not significantly predict changes in perceived social support, suggesting that the benefits of online peer support networks may be broadly accessible across personality profiles. Overall, the findings support the feasibility and psychological promise of structured digital peer support systems while highlighting the importance of cultural and personality-sensitive interpretations of mental health outcomes. Future research with larger samples and longer follow-up periods is warranted to clarify mechanisms, durability of effects, and optimal personalization strategies.

## Data Availability

Data cannot be shared publicly because of ethical restrictions regarding the privacy of human subjects and the sensitive nature of the data collected (e.g., mental health assessments and personal chat logs). De-identified, aggregated datasets necessary to replicate the study findings are available upon reasonable request to researchers who meet the criteria for access to confidential data. Requests for data access can be sent to the corresponding author.

